# SARS-CoV-2-specific T Cell Memory is Sustained in COVID-19 Convalescents for 8 Months with Successful Development of Stem Cell-like Memory T Cells

**DOI:** 10.1101/2021.03.04.21252658

**Authors:** Jae Hyung Jung, Min-Seok Rha, Moa Sa, Hee Kyoung Choi, Ji Hoon Jeon, Hyeri Seok, Dae Won Park, Su-Hyung Park, Hye Won Jeong, Won Suk Choi, Eui-Cheol Shin

**Author notes:** Corresponding authors Eui-Cheol Shin, M.D., Ph.D., Won Suk Choi, M.D., Ph.D., Hye Won Jeong, M.D., Ph.D. These authors contributed equally (Jae Hyung Jung, Min-Seok Rha).

## Abstract

Memory T cells contribute to rapid viral clearance during re-infection, but the longevity and differentiation of SARS-CoV-2-specific memory T cells remain unclear. We conducted direct *ex vivo* assays to evaluate SARS-CoV-2-specific CD4^+^ and CD8^+^ T cell responses in COVID-19 convalescents up to 254 days post-symptom onset (DPSO). Here, we report that memory T cell responses were maintained during the study period. In particular, we observed sustained polyfunctionality and proliferation capacity of SARS-CoV-2-specific T cells. Among SARS-CoV-2-specific CD4^+^ and CD8^+^ T cells detected by activation-induced markers, the proportion of stem cell-like memory T (T_SCM_) cells increased, peaking at approximately 120 DPSO. Development of T_SCM_ cells was confirmed by SARS-CoV-2-specific MHC-I multimer staining. Considering the self-renewal capacity and multipotency of T_SCM_ cells, our data suggest that SARS-CoV-2-specific T cells are long-lasting after recovery from COVID-19. The current study provides insight for establishing an effective vaccination program and epidemiological measurement.

## Introduction

Severe acute respiratory syndrome coronavirus 2 (SARS-CoV-2) infection causes coronavirus disease 2019 (COVID-19), an ongoing pandemic disease that threatens public health^1^. As of January 17, 2021, more than 93.2 million confirmed cases had been reported, and over 2 million deaths worldwide^2^. After SARS-CoV-2 infection, some patients, particularly elderly patients, develop severe COVID-19 that is associated with hyper- inflammatory responses^3,4^. Global efforts are underway to prevent the transmission of SARS-CoV-2 and to develop novel vaccines and therapeutic strategies. A thorough understanding of the immune responses against SARS-CoV-2 is urgently needed to control the COVID-19 pandemic.

Increasing evidence has demonstrated that SARS-CoV-2-specific memory T cell responses are elicited after recovery from COVID-19. A number of studies have reported SARS-CoV-2-specific memory T cell responses in the early convalescent phase of COVID-19^5-9^. SARS-CoV-2-specific CD4^+^ and CD8^+^ T cells have been detected in 100% and ∼70% of convalescent individuals a short time after resolution^5^. Recently, memory T cells were shown to contribute to protection against SARS-CoV-2 re-challenge in a rhesus macaque model^10^. Considering that T cell responses to SARS-CoV-1 and Middle East respiratory syndrome coronavirus (MERS-CoV) are long-lasting, up to >17 years^6,11-13^, SARS-CoV-2-specific memory T cells are expected to be maintained long-term and to contribute to rapid viral clearance during re-infection. A very recent study has examined SARS-CoV-2-specific T cell responses up to 8 months after infection using activation-induced marker (AIM) assays^14^.

Following natural infection or vaccination, the generation of effective and persistent T cell memory is essential for long-term protective immunity to the virus. Among diverse memory T cell subsets, stem cell-like memory T (T_SCM_) cells were recently reported to have the capacity for self-renewal and multipotency to repopulate the broad spectrum of memory and effector T cell subsets^15,16^. Thus, the successful generation of T_SCM_ cells is required for long-term protective T cell immunity^16^. For example, long-lived memory T cells following vaccination with live-attenuated yellow fever virus (YFV) exhibit stem cell-like properties and mediate lifelong protection^17,18^. However, limited knowledge is available on the differentiation of SARS-CoV-2-specific memory T cells following recovery from COVID-19, particularly the generation of T_SCM_ cells.

In the present study, we performed a comprehensive analysis of SARS-CoV-2-specific CD4^+^ and CD8^+^ T cell responses in peripheral blood mononuclear cells (PBMCs) from individuals with SARS-CoV-2 infection over 8 months post-infection. Using diverse T cell assays, we found that SARS-CoV-2-specific memory T cell responses were maintained 8 months after the infection. In addition, we analyzed the differentiation of SARS-CoV-2- specific memory T cells during the study period and revealed the successful generation of T_SCM_ cells. We also assessed the effector functions and proliferation capacity of long-term memory CD4^+^ and CD8^+^ T cells.

## Results

### Study cohort

We recruited 94 individuals with SARS-CoV-2 infection. The peak disease severity was evaluated according to the NIH severity of illness categories^19^: asymptomatic (n=6), mild (n=44), moderate (n=23), severe (n=13), and critical (n=8). Whole blood samples were obtained longitudinally (2-4 time points) from 38 patients or at a single time point from 56 patients. Whole blood was collected 1 to 254 days post-symptom onset (DPSO). Finally, a total of 160 PBMC samples were analyzed. Among 160 PBMC samples, 33 samples were obtained in the acute phase when viral RNA was still detected (1 - 33 DPSO), and 127 samples were obtained in the convalescent phase after the negative conversion of viral RNA (31 - 254 DPSO). In the current study, we defined the acute phase as 1 - 30 DPSO, and the convalescent phase as 31 - 254 DPSO. The demographic and clinical characteristics of enrolled patients are presented in Supplementary Table 1.

### SARS-CoV-2-specific T cell responses are sustained 8 months after the infection

First, we performed direct *ex vivo* interferon-γ (IFN-γ) enzyme-linked immunospot (ELISpot) assays following stimulation of PBMCs with overlapping peptide (OLP) pools spanning the spike (S), membrane (M), and nucleocapsid (N) proteins of SARS-CoV-2. S-, M-, and N-specific IFN-γ spot numbers increased during the acute phase. Subsequently, there was a decreasing tendency until 60 - 120 DPSO, and the IFN-γ responses were maintained over 8 months (Fig. 1a). These kinetics were also observed when S-, M-, and N-specific IFN-γ spot numbers were summed (Fig. 1a). In each PBMC sample, all three OLP pools evenly contributed to the IFN-γ responses without antigen dominance (Fig. 1b). We divided convalescent samples into three groups based on the DPSO at sample collection: T1 (31 - 99 DPSO), T2 (100 - 199 DPSO), and T3 (200 - 254 DPSO). We found no significant difference in IFN-γ spot numbers among groups (Fig. 1c), and an even contribution of antigens for the IFN- γ response was maintained among groups (Extended Data Fig. 1). Next, we focused on longitudinally tracked samples from 30 individuals in the convalescent phase. S-, M-, and N-specific and summed IFN-γ spot numbers were stable during the convalescent phase (Fig. 1d). When we compared paired samples from the same patient at two time points (t1, 31 - 100 DPSO; t2, 200 - 254 DPSO), we found no significant difference in IFN-γ spot numbers (Fig. 1e).

**Fig. 1.**
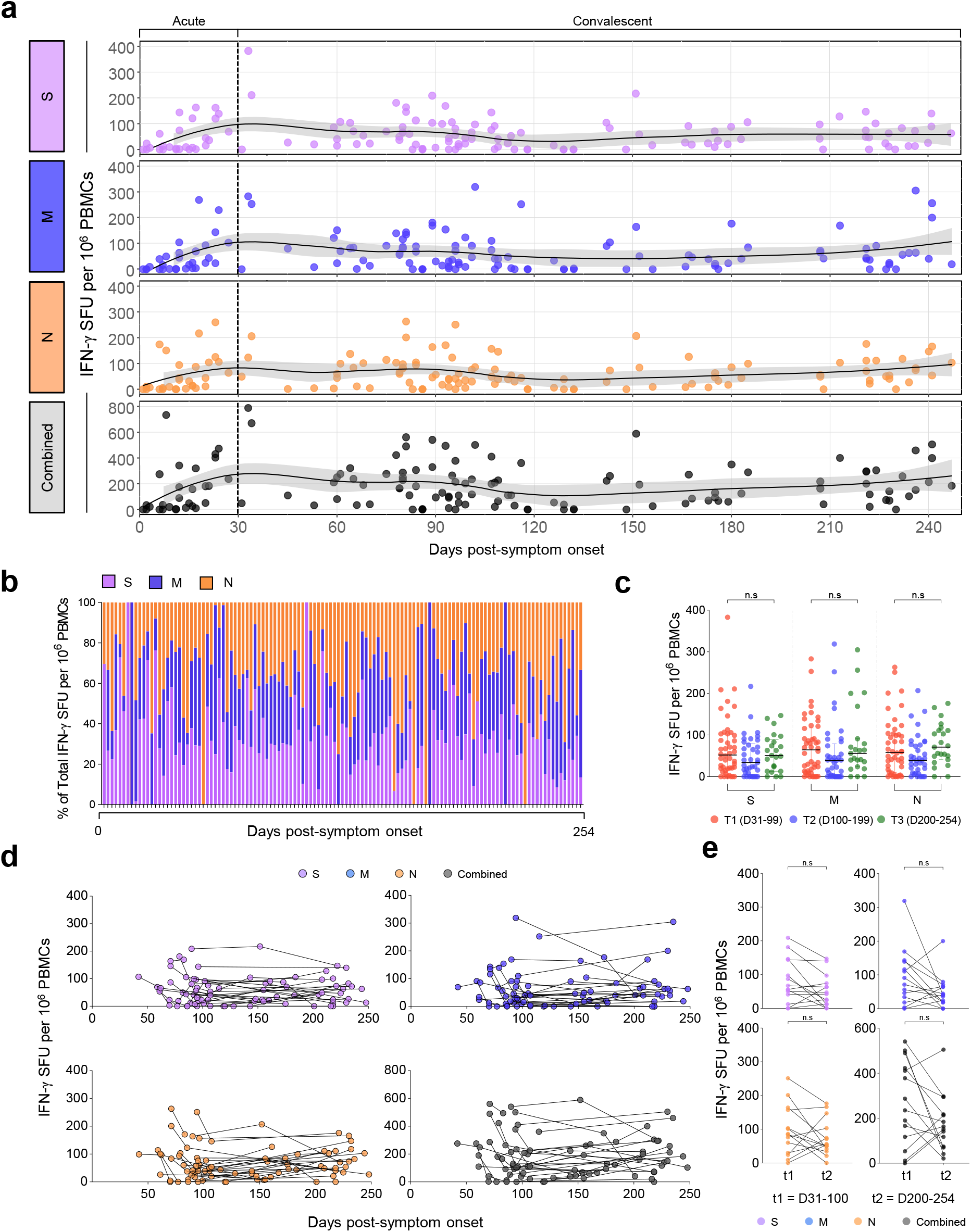
SARS-CoV-2-specific IFN-γ responses over 8 months post-infection. PBMC samples (n=135) from individuals with SARS-CoV-2 infection (n=76) were stimulated with OLPs of S, M, or N (1 μg/mL) for 24 h and the spot-forming units of IFN-γ-secreting cells examined by ELISpot. **a**, Scatter plots showing the relationship between DPSO and IFN-γ responses. The black line is a LOESS smooth nonparametric function, and the grey shading represents the 95% confidence interval. **b**, The composition of S-, M-, or N-specific IFN-γ responses among the total IFN-γ responses in each individual. **c**, IFN-γ responses were compared between T1 (n=46, 31 - 99 DPSO), T2 (n=37, 100 - 199 DPSO), and T3 (n=23, 200 - 254 DPSO). Data are presented as median and interquartile range (IQR). **d**,**e**, IFN-γ responses were analyzed in longitudinally tracked samples (n=82) from 30 individuals. **d**, Scatter plots showing the relationship between DPSO and IFN-γ responses. **e**, IFN-γ responses was compared between paired samples at two time points (n=14; t1, 31 - 100 DPSO; t2, 200 - 254 DPSO). Statistical analysis was performed using the Kruskal-Wallis test with Dunns’ multiple comparisons test (**c**) or the Wilcoxon signed-rank test (**e**). n.s, not significant.

We also evaluated SARS-CoV-2-specific T cell responses by AIM assays^5,20,21^ following stimulation of PBMCs with OLP pools of S, M, and N. CD137^+^OX40^+^ cells were considered SARS-CoV-2-specific cells among CD4^+^ T cells, and CD137^+^CD69^+^ cells were considered SARS-CoV-2-specific cells among CD8^+^ T cells^5^ (Fig. 2a, b). We confirmed a strong positive correlation between the frequency of CD137^+^OX40^+^ cells and the frequencies of alternative AIM^+^ cells (OX40^+^CD154^+^ or CD137^+^CD154^+^ cells) among CD4^+^ T cells (Extended Data Fig. 2). The frequency of S-, M-, and N-specific CD137^+^OX40^+^ cells among CD4^+^ T cells increased during the acute phase, decreased until 60 DPSO, and then was maintained over 8 months (Fig. 2c). When the frequency of S-, M-, and N-specific CD137^+^OX40^+^ cells was summed, similar kinetics were observed (Fig. 2c). The frequency of CD137^+^CD69^+^ cells among CD8^+^ T cells was relatively low compared to the frequency of CD137^+^OX40^+^ cells among CD4^+^ T cells, but exhibited similar kinetics (Fig. 2c). We also observed a positive correlation between the frequency of CD137^+^OX40^+^ cells among CD4^+^ T cells and the frequency of CD137^+^CD69^+^ cells among CD8^+^ T cells (Extended Data Fig. 3).

**Fig. 2.**
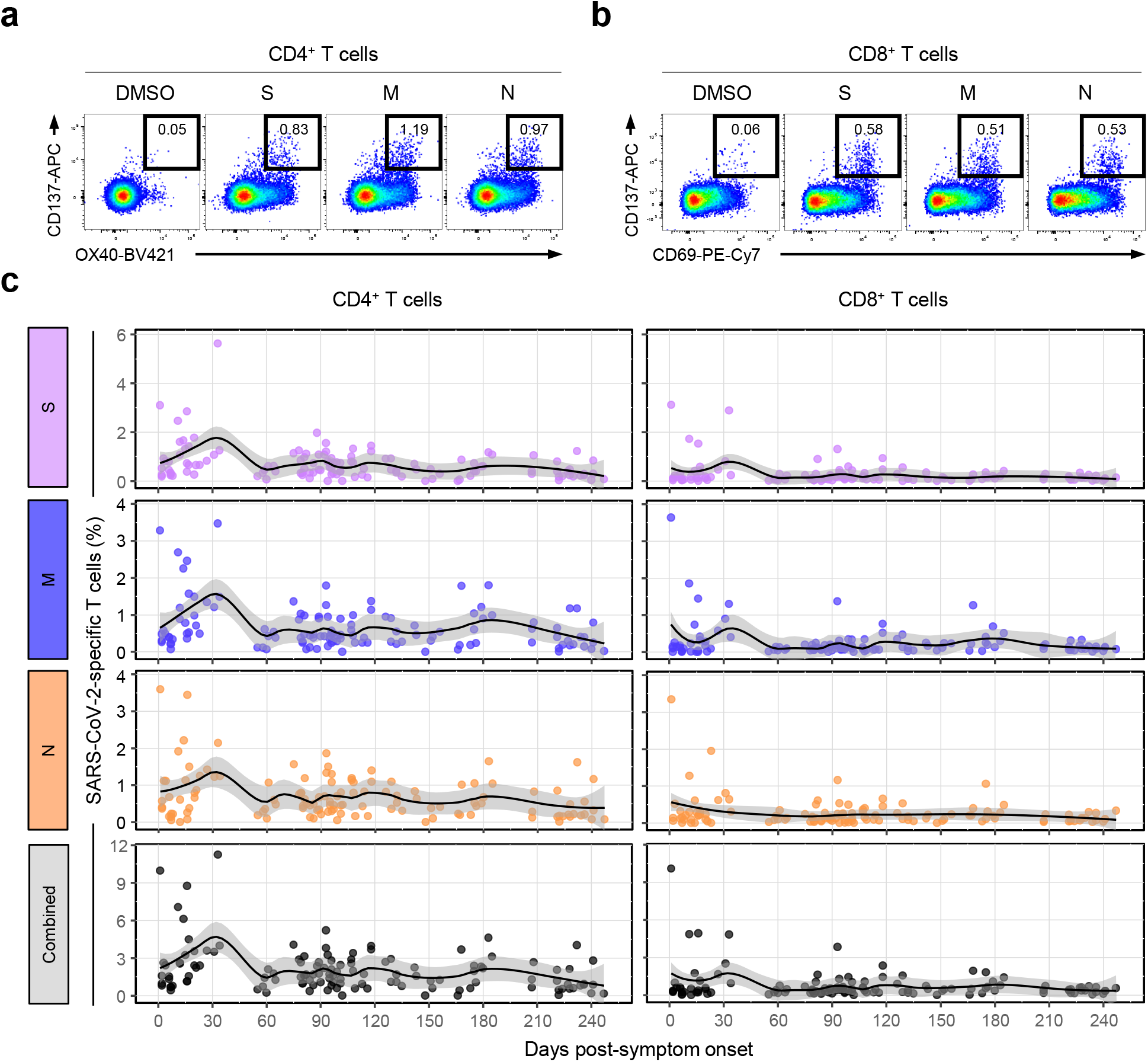
Kinetics of SARS-CoV-2-specific activation-induced marker (AIM)^+^ T cells. PBMC samples (n=125) from individuals with SARS-CoV-2 infection (n=69) were stimulated with OLPs of S, M, or N (1 μg/mL) for 24 h. The frequency of AIM^+^ (CD137^+^OX40^+^) cells among CD4^+^ T cells and the frequency of AIM^+^ (CD137^+^CD69^+^) cells among CD8^+^ T cells were analyzed. **a**,**b**, Representative flow cytometry plots showing the frequency of AIM^+^ cells among CD4^+^ (**a**) or CD8^+^ (**b**) T cells. **c**, Scatter plots showing the relationship between DPSO and the frequency of AIM^+^ cells among CD4^+^ (left) or CD8^+^ (right) T cells. The black is a LOESS smooth nonparametric function, and the grey shading represents the 95% confidence interval.

Collectively, these results indicate that SARS-CoV-2-specific T cell responses are long-lasting over 8 months in COVID-19 convalescents.

### SARS-CoV-2-specific T_SCM_ cells develop after the infection

Next, we examined the differentiation status of SARS-CoV-2-specific AIM^+^ T cells based on CCR7 and CD45RA expression in CD4^+^ (Fig. 3a) and CD8^+^ (Fig. 3b) T cell populations. Among SARS-CoV-2-specific AIM^+^CD4^+^ T cells, the proportion of CCR7^+^CD45RA^-^ (T_CM_) cells was maintained at approximately 50% on average during the study period, and the proportion of CCR7^-^CD45RA^-^ (T_EM_) cells increased up to approximately 35% on average until 60 DPSO and then maintained thereafter (Fig. 3c). CCR7^-^CD45RA^+^ (T_EMRA_) cells were a minor population (< 5%; Fig. 3c). Notably, the proportion of CCR7^+^CD45RA^+^ cells, which include both naïve and T_SCM_ cells, was approximately 30% on average in the acute phase, decreasing to approximately 10% on average by 60 DPSO and then maintained thereafter (Fig. 3c).

**Fig. 3.**
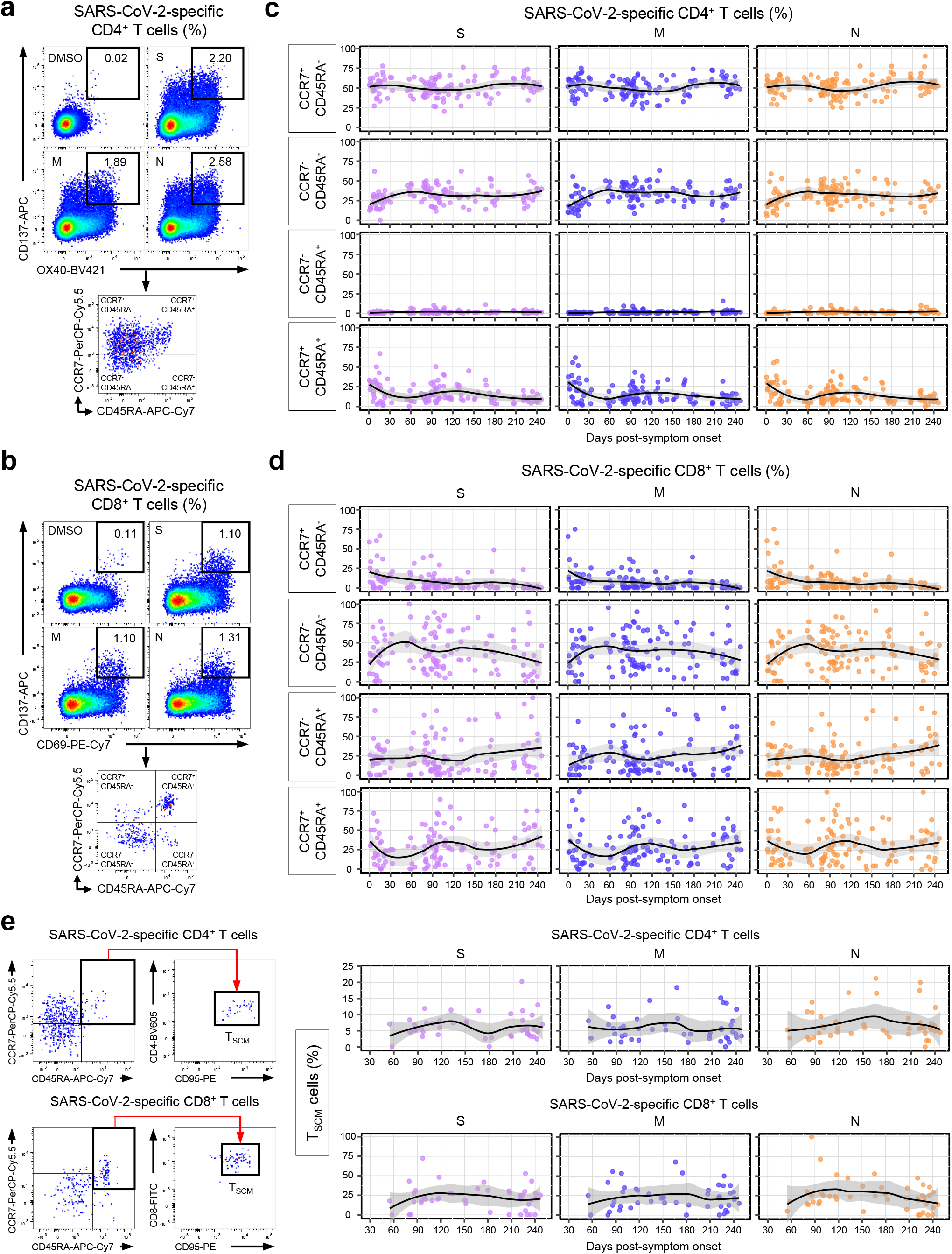
Differentiation status of SARS-CoV-2-specific AIM^+^ T cells. a-d, PBMC samples (n=124) from individuals with SARS-CoV-2 infection (n=68) were stimulated with OLPs of S, M, or N (1 μg/mL) for 24 h and the expression of CCR7 and CD45RA was analyzed in AIM^+^ (CD137^+^OX40^+^) CD4^+^ (**a**,**c**) and AIM^+^ (CD137^+^CD69^+^) CD8^+^ (**b**,**d**) T cells. **a**,**b**, Gating strategies for identifying each memory subset among AIM^+^CD4^+^ (**a**) or AIM^+^CD8^+^ (**b**) T cells. **c**,**d**, Scatter plots showing the relationship between DPSO and the proportion of the indicated subsets among AIM^+^CD4^+^ (**c**) or AIM^+^CD8^+^ (**d**) T cells. **e**, PBMC samples (n=47) from individuals with SARS-CoV-2 infection (n=34) were stimulated with OLPs of S, M, or N (1 μg/mL) for 24 h and the frequency of T_SCM_ (CCR7^+^CD45RA^+^CD95^+^) cells was analyzed in AIM^+^CD4^+^ (upper) and AIM^+^CD8^+^ (lower) T cells. Left, The gating strategy for identifying T_SCM_ cells. Right, Scatter plots showing the relationship between DPSO and the proportion of T_SCM_ cells among AIM^+^CD4^+^ or AIM^+^CD8^+^ T cells. The black line is a LOESS smooth nonparametric function, and the grey shading represents the 95% confidence interval (**c**,**d**,**e**).

Among SARS-CoV-2-specific AIM^+^CD8^+^ T cells, the proportion of CCR7^+^CD45RA^-^ (T_CM_) cells was approximately 20% on average in the acute phase and gradually decreased during the study period (Fig. 3d). The proportion of CCR7^-^CD45RA^-^ (T_EM_) cells increased up to 50% on average until 60 DPSO and was maintained thereafter (Fig. 3d). The proportion of CCR7^-^CD45RA^+^ (T_EMRA_) cells and CCR7^+^CD45RA^+^ cells was maintained (∼25% and 25-35% on average, respectively) during the study period (Fig. 3d).

We further investigated whether CCR7^+^CD45RA^+^ cells include T_SCM_ cells, which have a self-renewal capacity and multipotency for differentiation into diverse T cell subsets, by examining CD95, a marker of T_SCM_ cells^15,16^. In both AIM^+^CD4^+^ and AIM^+^CD8^+^ T cells, CD95^+^ cells were a dominant population among CCR7^+^CD45RA^+^ cells (Fig. 3e), indicating that the CCR7^+^CD45RA^+^ cells among AIM^+^CD4^+^ or AIM^+^CD8^+^ T cells are mainly T_SCM_ cells. The frequency of T_SCM_ cells among both AIM^+^CD4^+^ and AIM^+^CD8^+^ T cells gradually increased until 120 DPSO, when it reached a stable plateau (Fig. 3e). We found no significant difference in the frequency of T_SCM_ cells among AIM^+^CD4^+^ and AIM^+^CD8^+^ T cells between T1 (31 – 99 DPSO), T2 (100 - 199 DPSO), and T3 (200 - 254 DPSO; Extended Data Fig. 4a, b).

**Fig. 4.**
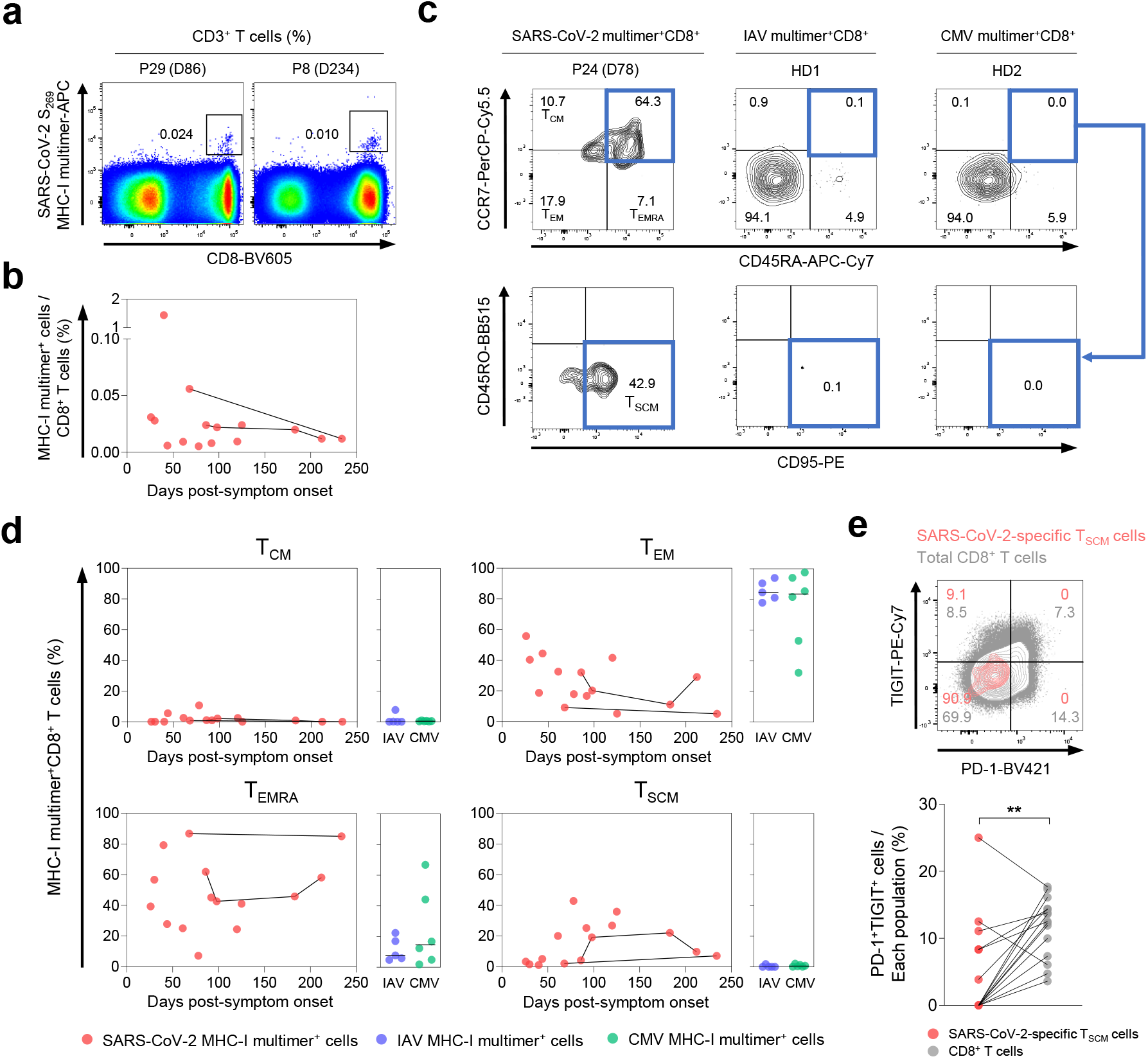
Frequency and differentiation status of SARS-CoV-2-specific MHC-I multimer^+^ T cells. PBMC samples (n=15) from individuals with SARS-CoV-2 infection (n=11) were analyzed by flow cytometry. **a**, Representative flow cytometry plots showing the *ex vivo* detection of SARS-CoV-2 S_269_ multimer^+^CD8^+^ T cells in the gate of CD3^+^ T cells. **b**, Scatter plot showing the relationship between DPSO and the frequency of SARS-CoV-2 S_269_ multimer^+^ cells among total CD8^+^ T cells. Samples from the same patient are connected by solid lines. **c**,**d**, The expression of CCR7, CD45RA, and CD95 was analyzed in SARS-CoV-2 S_269_ multimer^+^CD8^+^ T cells. IAV MP_58_ multimer^+^ (n=5) and CMV pp65_495_ multimer^+^ (n=6) cells from the PBMCs of healthy donors were also analyzed. Representative flow cytometry plots (**c**) show the proportion of the indicated subsets among multimer^+^ cells, and scatter plots (**d**) show the relationship between DPSO and the proportion of the indicated subsets among SARS-CoV-2 S_269_ multimer^+^ cells. Samples from the same patient are connected by solid lines. Summary data showing the proportion of the indicated subsets among IAV multimer^+^ and CMV multimer^+^ cells are also presented (**d**). Horizontal lines represent median. **e**, A representative flow cytometry plot (upper) and summary data (lower) showing the percentage of PD- 1^+^TIGIT^+^ cells among SARS-CoV-2 S_269_ multimer^+^ cells and total CD8^+^ T cells. Statistical analysis was performed using the Wilcoxon signed-rank test (**e**). **P < 0.01.

### Successful development of SARS-CoV-2-specific T_SCM_ cells is confirmed by direct ex vivo MHC-I multimer staining

To validate the results from *in vitro* stimulation-based AIM assays, we detected SARS-CoV-2-specific CD8^+^ T cells by performing direct *ex vivo* MHC-I multimer staining and examined the differentiation status of MHC-I multimer^+^ cells. We used an HLA-A*02 multimer loaded with SARS-CoV-2 S_269_ (YLQPRTFLL) peptide that has a low degree of homology to the human common cold coronaviruses (ccCoVs)^22,23^. MHC-I multimer^+^ cells were detected during the study period until 234 DPSO (Fig. 4a, b), and we determined the proportions of T_CM_ (CCR7^+^CD45RA^-^), T_EM_ (CCR7^-^CD45RA^-^), T_EMRA_ (CCR7^-^CD45RA^+^), and T_SCM_(CCR7^+^CD45RA^+^CD95^+^) cells among MHC-I multimer^+^ cells (Fig. 4c). Influenza A virus (IAV)- and cytomegalovirus (CMV)-specific MHC-I multimers were also used to stain PBMCs from healthy donors. Similar to the data from AIM^+^CD8^+^ T cells, T_EM_ and T_EMRA_ cells were the dominant populations among SARS-CoV-2-specific MHC-I multimer^+^ cells, whereas T_CM_ cells were a minor population during the study period (Fig. 4d). Among IAV- and CMV-specific MHC-I multimer^+^ cells from healthy donors, T_EM_ and T_EMRA_ cells were dominantly present, whereas T_CM_ cells were scarcely detected. T_SCM_ cells were also detected among SARS-CoV-2- specific MHC-I multimer^+^ cells during the study period, but not among IAV- and CMV-specific cells (Fig. 4d). In particular, the frequency of T_SCM_ cells among SARS-CoV-2-specific MHC-I multimer^+^ cells was high 60 – 125 DPSO.

A recent study has revealed two distinct subsets of CCR7^+^ stem cell-like progenitors: CCR7^+^PD-1^-^TIGIT^-^ cells with stem cell-like features and CCR7^+^PD-1^+^TIGIT^+^ cells with exhausted traits^24^. Therefore, we examined the expression of PD-1 and TIGIT in SARS-CoV-2-specific MHC-I multimer^+^ T_SCM_ cells. The frequency of PD-1^+^TIGIT^+^ cells was significantly lower among SARS-CoV-2-specific T_SCM_ cells than among total CD8^+^ T cells (Fig. 4e), confirming that SARS-CoV-2-specific T_SCM_ cells are *bona fide* stem-like memory cells.

### Polyfunctionality and proliferation capacity are preserved in long-term SARS-CoV-2- specific T cells

As we observed the generation of SARS-CoV-2-specific T_SCM_ cells with self-renewal capacity and multipotency, we aimed to examine the kinetics of the polyfunctionality of SARS-CoV-2-specific T cells during the study period. To this end, we performed intracellular cytokine staining of IFN-γ, IL-2, TNF, and CD107a following stimulation with OLP pools of S, M, and N and analyzed the polyfunctionality of CD4^+^ and CD8^+^ T cells (Fig. 5a). We defined polyfunctional cells as cells exhibiting positivity for ≥ 2 effector functions. Among SARS-CoV- 2-specific CD4^+^ and CD8^+^ T cells, the average proportion of polyfunctional cells was 25 - 40% and 30 - 50%, respectively, 60 DPSO, and was maintained until 254 DPSO (Fig. 5b). We found no significant difference in the frequency of polyfunctional cells among SARS-CoV-2-specific CD4^+^ and CD8^+^ T cells between T1 (31 - 99 DPSO), T2 (100 - 199 DPSO), and T3 (200 – 254 DPSO; Fig. 5c). Preserved polyfunctionality among SARS-CoV-2-specific CD4^+^ and CD8^+^ T cells was also observed when polyfunctionality was evaluated according to the number of positive effector functions (Fig. 5d).

**Fig. 5.**
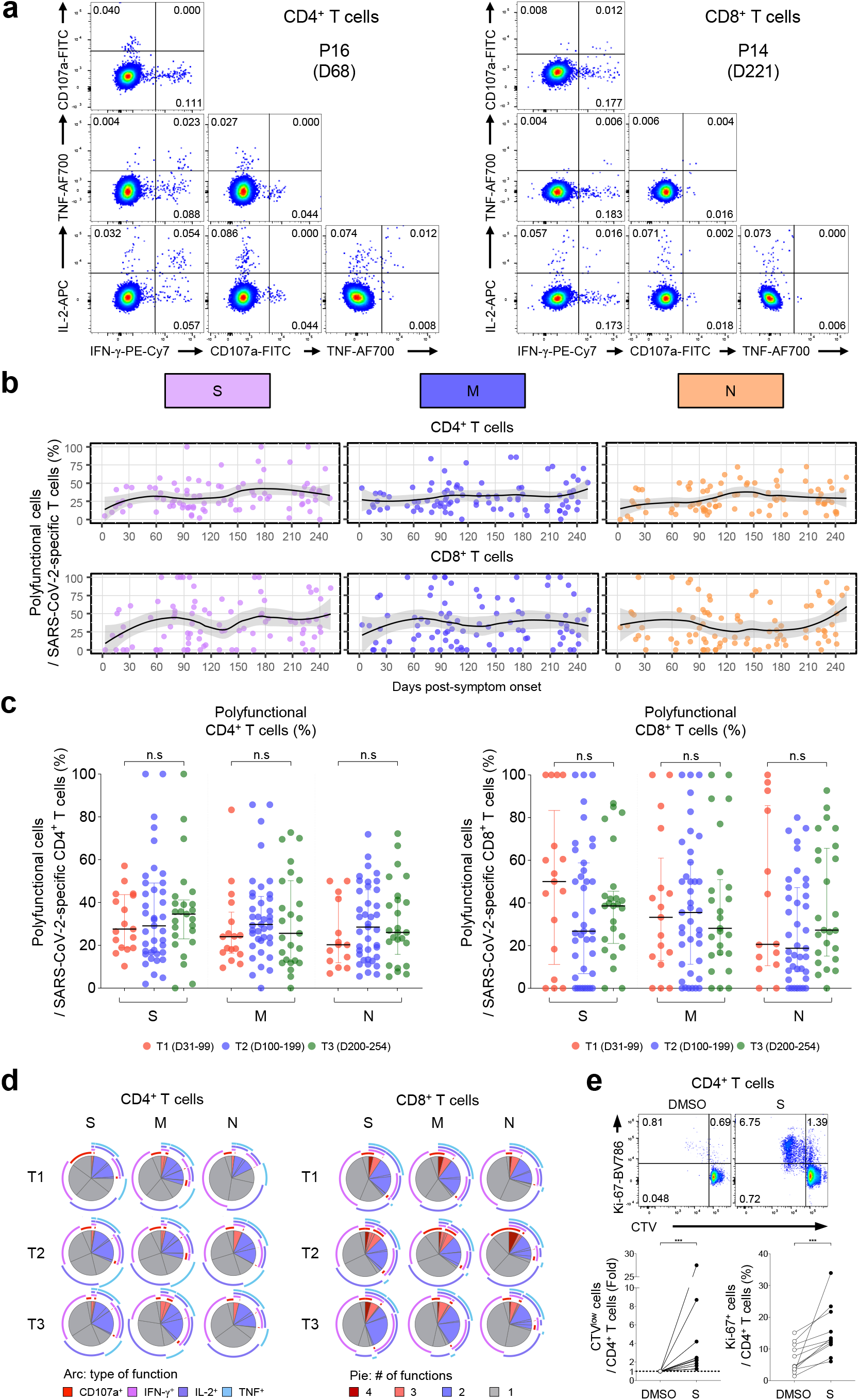
Polyfunctionality and proliferation capacity of SARS-CoV-2-specific T cells. a-d, PBMC samples (n=90) from individuals with SARS-CoV-2 infection (n=39) were stimulated with OLPs of S, M, or N (1 μg/mL) for 6 h. Intracellular cytokine staining was performed to examine the frequency of polyfunctional cells exhibiting positivity for ≥ 2 effector functions among SARS-CoV-2-specific CD4^+^ and CD8^+^ T cells. **a**, Representative flow cytometry plots showing the frequency of polyfunctional cells among CD4^+^ (left) and CD8^+^ (right) T cells. **b**, Scatter plots showing the relationship between DPSO and the frequency of polyfunctional cells among SARS-CoV-2-specific CD4^+^ (upper) or CD8^+^ (lower) T cells. The black line is a LOESS smooth nonparametric function, and the grey shading represents the 95% confidence interval. **c**, The fraction of polyfunctional cells among SARS-CoV-2-specific CD4^+^ (left) or CD8^+^ (right) T cells was compared between T1 (n=17, 31 - 99 DPSO), T2 (n=39, 100 - 199 DPSO), and T3 (n=25, 200 - 254 DPSO). Data are presented as median and IQR. **d**, Pie charts showing the fraction of cells positive for a given number of functions among SARS-CoV-2-specific CD4^+^ (left) or CD8^+^ (right) T cells. Each arc in the pie chart represents the indicated function. **e**, CTV-labeled PBMCs (n=11) obtained after 200 DPSO were stimulated with S OLP pool (1 μg/mL) for 120 h and the frequency of CTV^low^ and Ki-67^+^ cells among CD4^+^ T cells was analyzed. Representative plots (upper) and summary data (lower) are presented. Statistical analysis was performed using the Kruskal-Wallis test with Dunns’ multiple comparisons test (**c**) or the Wilcoxon signed-rank test (**e**). n.s, not significant, ***P < 0.001.

Finally, we examined the antigen-induced proliferation capacity of long-term SARS-CoV-2-specific memory T cells. We performed CellTrace Violet (CTV) dilution assays and Ki- 67 staining using PBMCs obtained after 200 DPSO. CD4^+^ T cells exhibited a robust proliferative response following *in vitro* stimulation with the S OLP pool (Fig. 5e), indicating that SARS-CoV-2-specific memory T cells elicit rapid recall responses upon viral re-exposure.

Considering preserved polyfunctionality and proliferation capacity of SARS-CoV-2- specific memory T cells, the current data indicate that memory T cells contribute to protective immunity against re-infection even 8 months after the primary infection.

## Discussion

Memory T cells play a crucial role in viral clearance during re-infection, but the longevity and differentiation status of SARS-CoV-2-specific memory T cells among COVID-19 convalescents remain unclear. In the present study, we demonstrated that SARS-CoV-2- specific memory T cell responses were maintained in COVID-19 convalescents 8 months post-infection. Notably, we found that SARS-CoV-2-specific T_SCM_ cells were successfully developed, indicating that SARS-CoV-2-specific T cell memory may be long-lasting in COVID- 19 convalescents. These findings were supported by the SARS-CoV-2-specific T cells from PBMCs obtained after 200 DPSO exhibiting sustained polyfunctionality and proliferation capacity. Our results may fill a gap in understanding T cell memory responses after recovery from SARS-CoV-2 infection.

Recent studies suggest critical roles of T cells in the clearance of SARS-CoV-2 and protection from developing severe COVID-19. In particular, it has been shown that coordination of adaptive immune responses, including CD4^+^ T cell, CD8^+^ T cell, and antibody responses, is essential for controlling COVID-19^20^. Notably, peak disease severity has been shown to inversely correlate with the frequency of SARS-CoV-2-specific CD4^+^ and CD8^+^ T cells instead of SARS-CoV-2 antibody titers^20^. In another study, T cell responses were detected among COVID-19 convalescents without detectable SARS-CoV-2 IgG^25^. In a rhesus macaque model, CD8-depleted convalescent animals exhibited limited viral clearance in the respiratory tract upon SARS-CoV-2 re-challenge, suggesting that CD8^+^ T cells contribute to viral clearance during SARS-CoV-2 re-infection^10^. Collectively, these studies strongly demonstrate a protective role of T cells in COVID-19.

Currently, we do not have exact information on how long adaptive immune memory lasts in COVID-19 convalescents. Whether antibody responses to SARS-CoV-2 wane over time in patients who have recovered from COVID-19 remains controversial^26-28^. Previous studies on SARS-CoV-1 and MERS-CoV infection have shown that T cell responses were more enduring compared to antibody responses^12,13^. A recent study detected SARS-CoV-1-specific T cell responses 17 years after infection^6^. Similarly, slow decay of SARS-CoV-2-specific memory T cells is expected. In the present study, we demonstrated the maintenance of SARS-CoV-2-specific memory T cell responses in COVID-19 convalescents over 8 months post-infection using a battery of T cell assays. Given that vaccination programs for prophylaxis of SARS-CoV-2 infection are being launched worldwide, another question is how long memory T cells elicited by vaccination will last. Vaccine-induced memory T cells may differ in phenotype and durability from infection-induced memory T cells, which should be addressed in future studies.

A series of studies have reported the existence of SARS-CoV-2-reactive T cell responses among unexposed individuals, suggesting that memory T cells induced by previous ccCoV infection are cross-reactive to SARS-CoV-2 proteins^5,29-32^. Therefore, in the present study, we could not distinguish *de novo* primed T cells by SARS-CoV-2 infection from pre-existing, cross-reactive ccCoV-specific T cells. However, we could selectively examine *de novo* primed SARS-CoV-2-specific T cells by staining with an HLA-A*02 multimer loaded with SARS-CoV-2 S_269_ (YLQPRTFLL) peptide. This epitope peptide has a low degree of homology with ccCoVs, including OC43, HKU1, 229E, and NL63^22,23^. In future studies of SARS-CoV-2- specific T cell responses, cross-reactive epitope peptides and SARS-CoV-2-specific epitope peptides will need to be used separately^30^.

Among distinct subsets of memory T cells, T_SCM_ cells possess a superior ability for self-renewal, memory recall responses, and multipotency to reconstitute diverse memory subsets^15^. Therefore, long-term T cell memory relies on the successful generation of T_SCM_ cells^16^. Previous studies showed that long-lasting YFV-specific CD8^+^ T cells resemble T_SCM_ cells^17,18^. We and others have also suggested the generation of SARS-CoV-2-specific T_SCM_ cells in the convalescent phase of COVID-19 on the basis of the expression of CCR7 and CD45RA^8,23^. However, these studies on COVID-19 patients did not examine the definitive marker of T_SCM_ cells, CD95. In the present study, we delineated the kinetics of T_SCM_ cells using both AIM assays and MHC-I multimer staining, and observed the successful generation of T_SCM_ cells in COVID-19 convalescents. In line with these findings, we also found sustained polyfunctionality and proliferation capacity, suggesting efficient memory recall responses. A recent study has proposed that CCR7^+^ stem-like progenitor cells are composed of two separate populations, which are distinguished by PD-1 and TIGIT expression^24^. We demonstrated that PD-1 and TIGIT are rarely expressed in SARS-CoV-2-specific T_SCM_ cells, indicating that SARS-CoV-2-specific T_SCM_ cells are not exhausted-like progenitors, but *bona fide* stem-like memory cells.

Polyfunctional T cells, which exert multiple effector functions simultaneously, play a critical role in host protection against viral infection^33-35^. For example, polyfunctional CD8^+^ T cells are preserved in human immunodeficiency virus-infected long-term non-progressors^34^. In addition, polyfunctional T cell responses are associated with effective control of hepatitis C virus (HCV)^35^. It has also been reported that virus-specific polyfunctional T cells can be successfully developed by immunization with vaccinia virus^36^ and an HCV vaccine^37^. Collectively, polyfunctional memory T cells control viral infection more efficiently than monofunctional T cells. Therefore, generation of polyfunctional memory T cells following natural infection or vaccination is expected to confer protective immunity. In this regard, the sustained polyfunctionality of long-term SARS-CoV-2-specific T cells observed in our study is highly suggestive of long-lasting protective immunity in COVID-19 convalescents.

In summary, we conducted a comprehensive analysis of SARS-CoV-2-specific memory T cell responses over 8 months post-infection. Our current analysis provides valuable information regarding the longevity and differentiation of SARS-CoV-2-specific memory T cells elicited by natural infection. These data add to our basic understanding of memory T cell responses in COVID-19, which aids in establishing an effective vaccination program and epidemiological measurement.

## Methods

### Patients and specimens

In this study, 94 patients with PCR-confirmed SARS-CoV-2 infection were enrolled from Ansan Hospital and Chungbuk National University Hospital, Republic of Korea. Peripheral blood was obtained from all patients with SARS-CoV-2 infection. We also obtained peripheral blood from seven healthy donors. In six asymptomatic patients, the date of the first admission was regarded as 7 DPSO because the date of the first admission among symptomatic patients was an average 7 DPSO. This study was reviewed and approved by the institutional review board of all participating institutions and conducted according to the principles of the Declaration of Helsinki. Informed consent was obtained from all donors and patients.

PBMCs were isolated by density gradient centrifugation using Lymphocyte Separation Medium (Corning). After isolation, the cells were cryopreserved in fetal bovine serum (FBS; Corning) with 10% dimethyl sulfoxide (DMSO; Sigma-Aldrich) until use.

### Ex vivo IFN-γ enzyme-linked immunospot assay

Plates with hydrophobic polyvinylidene difluoride membrane (Millipore, Billerica, MA) were coated with 2 μg/mL anti-human monoclonal IFN-γ coating antibody (clone 1-D1K, Mabtech) overnight at 4°C. The plates were washed with sterile phosphate-buffered saline (PBS) and blocked with 1% bovine serum albumin (Bovogen) for 1 hour at room temperature (RT). 700,000 PBMCs were seeded per well and stimulated with 1 μg/mL OLP pools spanning SARS-CoV-2 spike, membrane, and nucleocapsid proteins (Miltenyi Biotec) for 24 hours at 37°C. We used 10 μg/mL phytohemagglutinin as a positive control and an equimolar amount of DMSO as a negative control. Plates were washed with 0.05% Tween-PBS (Junsei Chemical) and incubated with 0.25 μg/mL biotinylated anti-human monoclonal IFN-γ antibody (clone 7-B6-1, Mabtech) for 2 hours at RT. After washing, streptavidin-alkaline phosphatase (Invitrogen) was added sequentially. Precipitates were detected with AP color reagent (Bio-Rad) and the reaction stopped by rinsing with distilled water. Spot-forming units were quantified using an automated ELISpot reader (AID). To quantify SARS-CoV-2-specific responses, spots in the negative control wells were subtracted from the OLP-stimulated wells.

### Multi-color flow cytometry

Cells were stained with fluorochrome-conjugated antibodies for specific surface markers for 10 minutes at RT. Dead cells were excluded using LIVE/DEAD red fluorescent reactive dye (Invitrogen). In intracellular staining experiments, cells were fixed and permeabilized using the FoxP3 staining buffer kit (Invitrogen), and then stained for intracellular markers for 30 minutes at 4°C. Multi-color flow cytometry was performed using an LSR II instrument (BD Biosciences) and the data analyzed in FlowJo software (FlowJo LLC). The fluorochrome-conjugated MHC-I multimers and monoclonal antibodies used in this study are listed in Supplementary Table 2.

### Activation-induced marker assay

PBMCs were blocked with 0.5 μg/mL anti-human CD40 mAb (clone HB14, Miltenyi Biotec) in RPMI 1640 supplemented with 10% FBS and 1% penicillin and streptomycin for 15 minutes at 37°C. The cells were then cultured in the presence of 1 μg/mL SARS-CoV-2 OLP pools and 1 μg/mL anti-human CD28 and CD49d mAbs (clone L293 and L25, respectively, BD Biosciences) for 24 hours. Stimulation with an equal concentration of DMSO in PBS was performed as a negative control.

### MHC-I multimer staining

PBMCs were stained with APC-conjugated MHC-I SARS-CoV-2 S_269_ pentamer (Proimmune), IAV MP_58_ dextramer (Immudex), or CMV pp65_495_ dextramer (Immudex) for 15 minutes at RT and washed twice. Additional surface markers were stained using the protocols described above.

### Stimulation for intracellular cytokine staining

PBMCs were cultured in the presence of SARS-SoV-2 OLP pools and 1 μg/mL anti-human CD28 and CD49d mAbs for 6 hours at 37°C. Brefeldin A (GolgiPlug, BD Biosciences) and monensin (GolgiStop, BD Biosciences) were added 1 hour after the initial stimulation.

### Proliferation assays

PBMCs were labeled using the CellTrace™ Violet Cell Proliferation Kit (Invitrogen) at a concentration of 1.0 × 10^6^ cells/mL for 20 minutes at 37°C. We then added 1% FBS-PBS to the cells for 5 minutes at RT to quench unbound dyes. Cells were washed and cultured in RPMI 1640 (Corning) containing 10% FBS (Corning) and 1% penicillin-streptomycin (Welgene) at a concentration of 500,000 cells per well in the presence of 1 μg/mL SARS-CoV- 2 spike peptide pools and 1 μg/mL anti-human CD28 and CD49d mAbs (clone L293 and L25 respectively, BD Biosciences) for 120 hours. Stimulation with an equal concentration of DMSO in PBS was performed as a negative control. After incubation, cells were harvested and stained with antibodies for analysis by flow cytometry.

### Data quantification and statistical analysis

Data were calculated as background-subtracted data. Background-subtracted data were derived by subtracting the value after DMSO stimulation. When three stimuli were combined, the value after each stimulation was combined and we subtracted triple the value derived from DMSO stimulation.

Statistical analyses were performed using GraphPad Prism version 9 for Windows (GraphPad Software). Significance was set at p<0.05. The Wilcoxon signed-rank test was used to compare data between two paired groups. In addition, the Kruskal-Wallis test with Dunns’ multiple comparisons test was used to compare non-parametric data between multiple unpaired groups. To assess the significance of correlation, the Spearman correlation test was used. In cross-sectional analyses, a non-parametric local regression (LOESS) function was employed with 95% confidence interval.

## Data Availability

The data that support the findings of this study are available from the corresponding author upon request.

## Data availability

Data relating to the findings of this study are available from the corresponding author upon reasonable request.

## Acknowledgments

This work was supported by the Samsung Science and Technology Foundation under Project Number SSTF-BA1402-51, and the Mobile Clinic Module Project funded by KAIST.

## Author Contributions

J.H.J., M.-S.R., H.W.J., W.S.C., and E.-C.S. designed the research. H.K.C., J.H.J., H.S., D.W.P., H.W.J., and W.S.C. collected clinical specimens. J.H.J., M.-S.R., and M.S. performed experiments. J.H.J., M.-S.R., S.-H.P., and E.-C.S. analyzed the results. J.H.J., M.-S.R. and E.- C.S. wrote the manuscript.

## Figure Legends

**Supplementary Table 1.** Characteristics of enrolled patients

| Parameter | COVID-19 (n=94) |
| --- | --- |
| Age(years) | 19-96 (median 39, IQR 29) |
| Gender (F/M) | 55/39 |
| Peak disease severity<sup>a</sup> |  |
| Asymptomatic, n | 6 (6.4%) |
| Mild, n | 44 (46.8%) |
| Moderate, n | 23 (24.5%) |
| Severe, n | 13 (13.8%) |
| Critical, n | 8 (8.5%) |
| DPSO at sample collection | 1-254 (median 94, IQR 93) |
| Blood collection |  |
| Multiple time points, n | 38 |
| 2 | 16 |
| 3 | 16 |
| 4 | 6 |
| Single time point, n | 56 |
| Assays | Total 160 samples |
| IFN-γ ELISpot assays | 135 samples (n=76) |
| ICS | 90 samples (n=39) |
| AIM assays | 125 samples (n=69) |
| MHC-I multimer staining | 15 samples (n=11) |
| Proliferation assays | 11 samples (n=11) |

IQR, interquartile range; DPSO, days post-symptom onset; ELISpot, enzyme-linked immunospot; ICS, intracellular cytokine staining; AIM, activation-induced marker; CTV, CellTrace Violet.
<sup>a</sup>Disease severity was defined by the NIH severity of illness categories.

**Supplementary Table 2.**
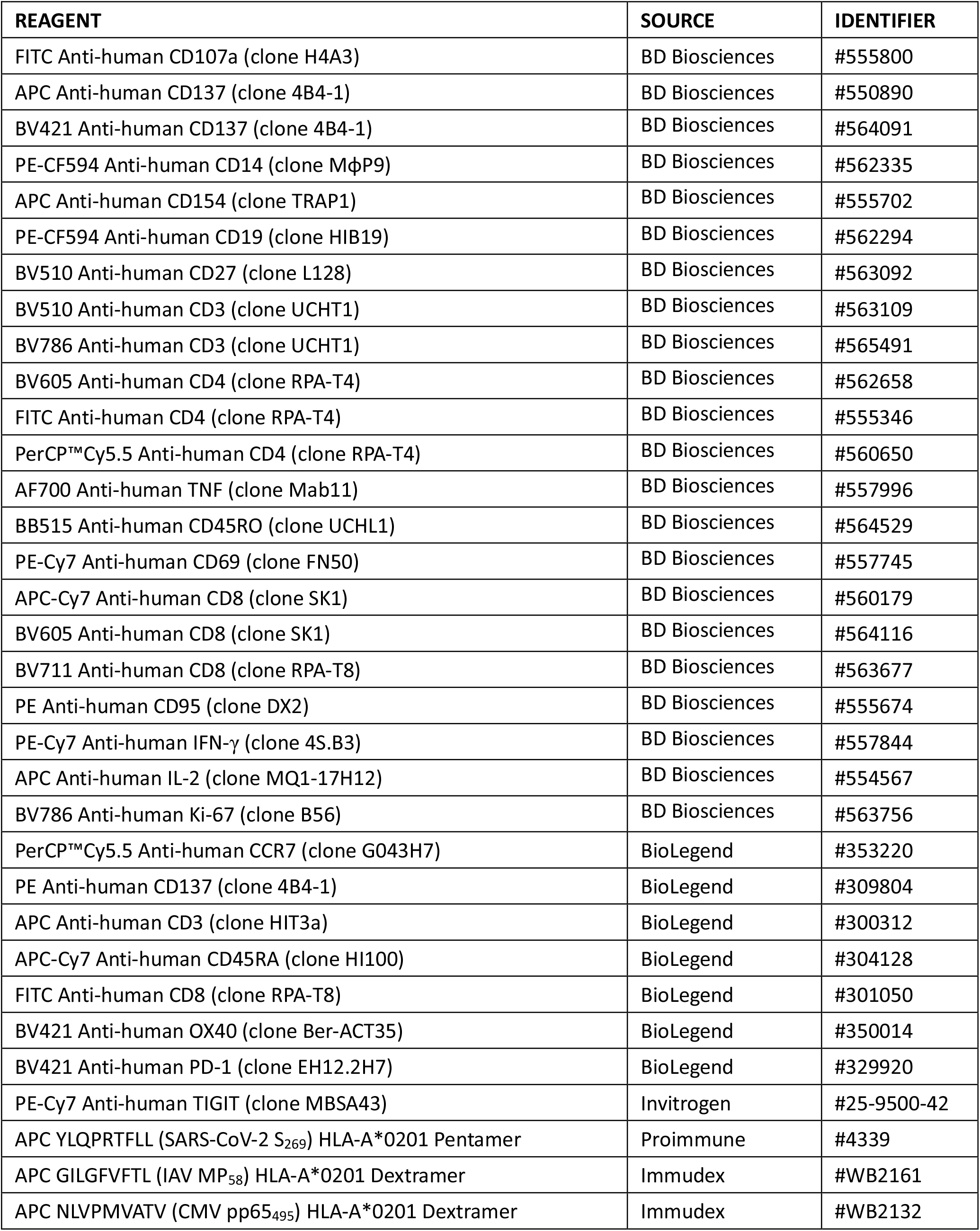
Flow cytometry reagents

**Extended Data Fig. 1.**
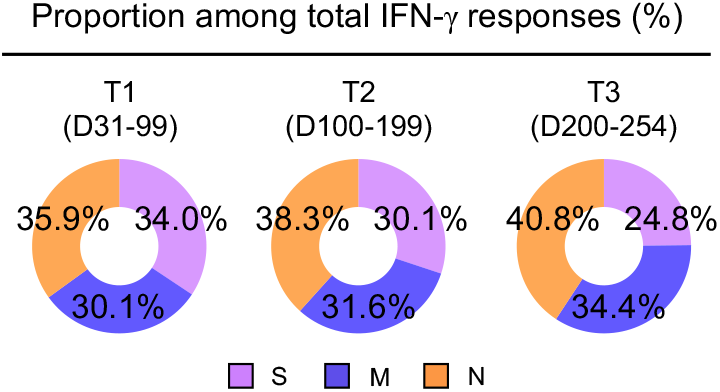
Proportion of S-, M-, and N-specific IFN-γ responses among total IFN-γ responses. PBMC samples from individuals with SARS-CoV-2 infection were stimulated with OLPs of S, M, or N (1 μg/mL) for 24 h and spot-forming units of IFN-γ-secreting cells were examined by ELISpot. Pie charts showing the proportion of S-, M-, and N-specific IFN-γ responses among the total IFN-γ responses in T1 (n=46, 31 - 99 DPSO), T2 (n=37, 100 - 199 DPSO), and T3 (n=23, 200 - 254 DPSO).

**Extended Data Fig 2.**
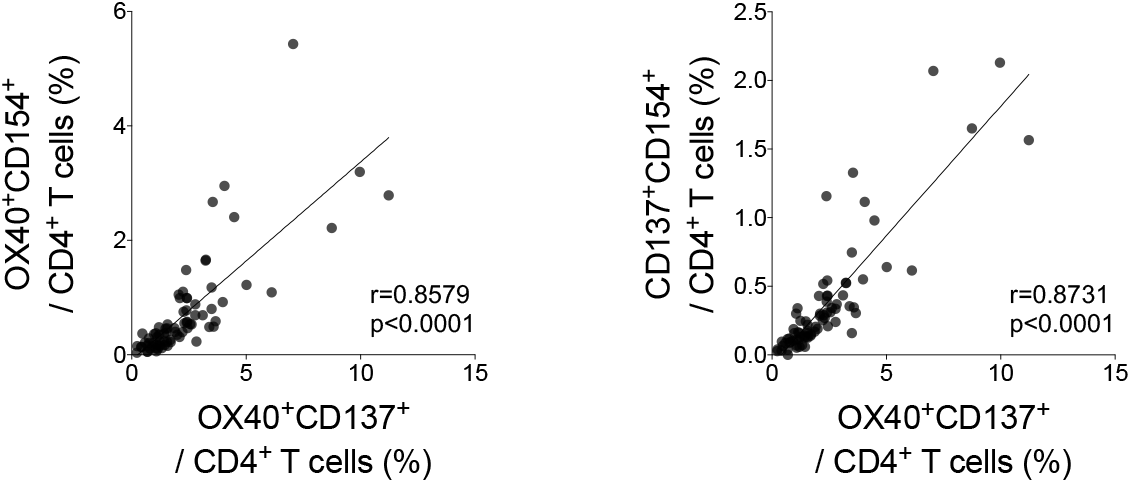
Correlation between the frequency of CD137^+^OX40^+^ cells and the frequencies of alternative AIM^+^ cells (OX40^+^CD154^+^ or CD137^+^CD154^+^ cells) among CD4^+^ T cells. PBMC samples from individuals with SARS- CoV-2 infection were stimulated with OLPs of S, M, or N (1 μg/mL) for 24 hours, and the correlation between the frequency of CD137^+^OX40^+^ cells and the frequencies of alternative AIM^+^cells (OX40CD154^+^ or CD137^+^CD154^+^ cells) among CD4^+^ T cells was analyzed (n=78). Statistical analysis was performed using the Spearman correlation test.

**Extended Data Fig. 3.**
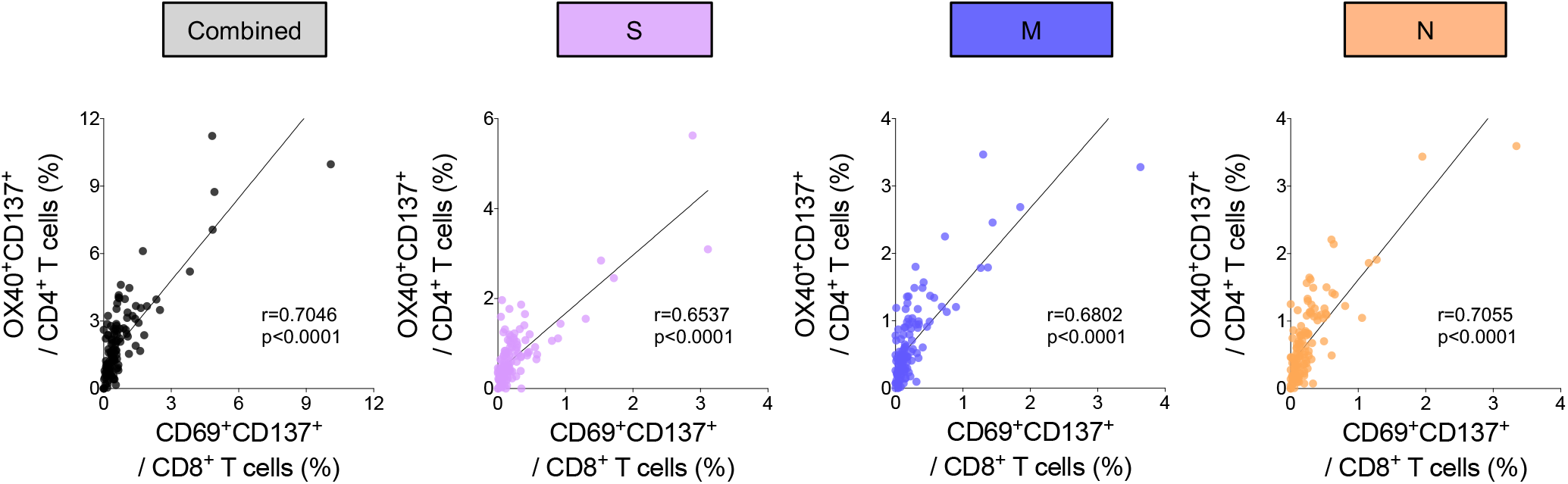
Correlation of the frequency of AIM^+^ cells between CD4^+^ and CD8^+^ T cells. PBMC samples from individuals with SARS-CoV-2 infection were stimulated with OLPs of S, M, or N (1 μg/mL) for 24 h and the correlation between the frequency of AIM^+^ (CD137^+^OX40^+^) cells among CD4^+^ T cells and AIM^+^ (CD137^+^CD69^+^) cells among CD8^+^ T cells was analyzed (n=125). Statistical analysis was performed using the Spearman correlation test.

**Extended Data Fig. 4.**
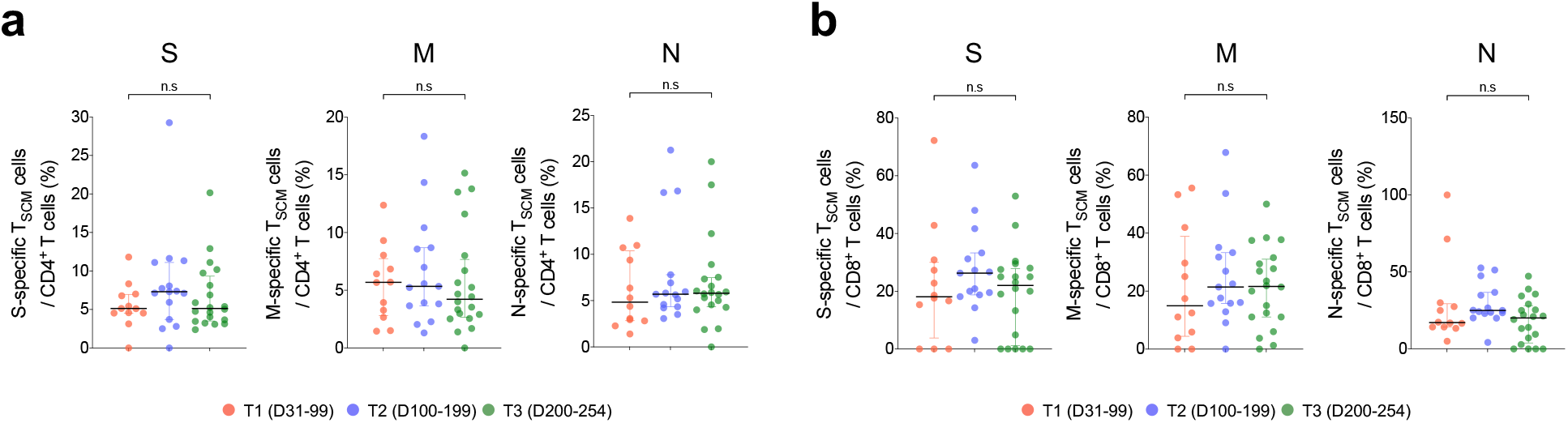
Frequency of T_SCM_ cells among SARS-CoV-2-specific T cells according to days post-symptom onset. a,b, PBMC samples from individuals with SARS-CoV-2 infection were stimulated with OLPs of S, M, or N (1 μg/mL) for 24 h and the frequency of T_SCM_ (CCR7^+^CD45RA^+^CD95^+^) cells was analyzed among AIM^+^ (CD137^+^OX40^+^) CD4^+^ (**a**) and AIM^+^ (CD137^+^CD69^+^) CD8^+^ T cells (**b**). The frequencies of T_SCM_ cells were compared between T1 (n=12, 31 - 99 DPSO), T2 (n=14, 100 - 199 DPSO), and T3 (n=20, 200 - 254 DPSO). Data are presented as median and IQR. Statistical analysis was performed using the Kruskal-Wallis test with Dunns’ multiple comparisons test. n.s, not significant.

## References

1 Huang, C. et al. Clinical features of patients infected with 2019 novel coronavirus in Wuhan, China. Lancet 395, 497–506 (2020).

2 World Health Organization. COVID-19 Weekly Epidemiological Update-19 January 2020 https://www.who.int/publications/m/item/weekly-epidemiological-update-19-january-2020 (2020).

3 Lee, J. S. et al. Immunophenotyping of COVID-19 and influenza highlights the role of type I interferons in development of severe COVID-19. Sci. Immunol. 5, eabd1554 (2020).

4 Lee, J. S. & Shin, E. C. The type I interferon response in COVID-19: implications for treatment. Nat. Rev. Immunol. 20, 585–586 (2020).

5 Grifoni, A. et al. Targets of T Cell Responses to SARS-CoV-2 Coronavirus in Humans with COVID-19 Disease and Unexposed Individuals. Cell 181, 1489-1501.e15 (2020).

6 Le Bert, N. et al. SARS-CoV-2-specific T cell immunity in cases of COVID-19 and SARS, and uninfected controls. Nature 584, 457–462 (2020).

7 Peng, Y. et al. Broad and strong memory CD4(+) and CD8(+) T cells induced by SARS-CoV-2 in UK convalescent individuals following COVID-19. Nat. Immunol. 21, 1336–1345 (2020).

8 Sekine, T. et al. Robust T Cell Immunity in Convalescent Individuals with Asymptomatic or Mild COVID-19. Cell 183, 158-168.e14 (2020).

9 Rodda, L. B. et al. Functional SARS-CoV-2-Specific Immune Memory Persists after Mild COVID-19. Cell 184, 169-183.e17 (2020).

10 McMahan, K. et al. Correlates of protection against SARS-CoV-2 in rhesus macaques. Nature, doi:10.1038/s41586-020-03041-6 (2020).

11 Ng, O. W. et al. Memory T cell responses targeting the SARS coronavirus persist up to 11 years post-infection. Vaccine 34, 2008–2014 (2016).

12 Zhao, J. X. et al. Recovery from the Middle East respiratory syndrome is associated with antibody and T cell responses. Sci. Immunol. 2, eaan5393 (2017).

13 Tang, F. et al. Lack of peripheral memory B cell responses in recovered patients with severe acute respiratory syndrome: a six-year follow-up study. J. Immunol. 186, 7264–7268 (2011).

14 Dan, J. M. et al. Immunological memory to SARS-CoV-2 assessed for up to 8 months after infection. Science, doi:10.1126/science.abf4063 (2021).

15 Gattinoni, L. et al. A human memory T cell subset with stem cell-like properties. Nat. Med. 17, 1290–1297 (2011).

16 Gattinoni, L., Speiser, D. E., Lichterfeld, M. & Bonini, C. T memory stem cells in health and disease. Nat. Med. 23, 18–27 (2017).

17 Fuertes Marraco, S. A. et al. Long-lasting stem cell-like memory CD8+ T cells with a naive-like profile upon yellow fever vaccination. Sci. Transl. Med. 7, 282ra248 (2015).

18 Akondy, R. S. et al. Origin and differentiation of human memory CD8 T cells after vaccination. Nature 552, 362–367 (2017).

19 National Institutes of Health. COVID-19 Treatment Guidelines Panel. Coronavirus Disease 2019 (COVID-19) Treatment Guidelines. https://www.covid19treatmentguidelines.nih.gov (2020).

20 Rydyznski Moderbacher, C. et al. Antigen-Specific Adaptive Immunity to SARS-CoV-2 in Acute COVID-19 and Associations with Age and Disease Severity. Cell 183, 996-1012.e19 (2020).

21 Morou, A. et al. Altered differentiation is central to HIV-specific CD4(+) T cell dysfunction in progressive disease. Nat. Immunol. 20, 1059–1070 (2019).

22 Shomuradova, A. S. et al. SARS-CoV-2 Epitopes Are Recognized by a Public and Diverse Repertoire of Human T Cell Receptors. Immunity 53, 1245-1257.e5 (2020).

23 Rha, M. S. et al. PD-1-Expressing SARS-CoV-2-Specific CD8(+) T Cells Are Not Exhausted, but Functional in Patients with COVID-19. Immunity 54, 44-52.e3 (2021).

24 Galletti, G. et al. Two subsets of stem-like CD8(+) memory T cell progenitors with distinct fate commitments in humans. Nat. Immunol. 21, 1552–1562 (2020).

25 Schwarzkopf, S. et al. Cellular Immunity in COVID-19 Convalescents with PCR-Confirmed Infection but with Undetectable SARS-CoV-2-Specific IgG. Emerg. Infect. Dis. 27, doi:10.3201/2701.203772 (2020).

26 Ibarrondo, F. J. et al. Rapid Decay of Anti-SARS-CoV-2 Antibodies in Persons with Mild Covid-19. N. Engl. J. Med. 383, 1085–1087 (2020).

27 Isho, B. et al. Persistence of serum and saliva antibody responses to SARS-CoV-2 spike antigens in COVID-19 patients. Sci. Immunol. 5, eabe5511 (2020).

28 Long, Q. X. et al. Clinical and immunological assessment of asymptomatic SARS-CoV-2 infections. Nat. Med. 26, 1200–1204 (2020).

29 Mateus, J. et al. Selective and cross-reactive SARS-CoV-2 T cell epitopes in unexposed humans. Science 370, 89–94 (2020).

30 Nelde, A. et al. SARS-CoV-2-derived peptides define heterologous and COVID-19-induced T cell recognition. Nat. Immunol. 22, 74–85 (2020).

31 Woldemeskel, B. A. et al. Healthy donor T cell responses to common cold coronaviruses and SARS-CoV-2. J. Clin. Invest. 130, 6631–6638 (2020).

32 Braun, J. et al. SARS-CoV-2-reactive T cells in healthy donors and patients with COVID-19. Nature 587, 270–274 (2020).

33 Almeida, J. R. et al. Superior control of HIV-1 replication by CD8+ T cells is reflected by their avidity, polyfunctionality, and clonal turnover. J. Exp. Med. 204, 2473–2485 (2007).

34 Betts, M. R. et al. HIV nonprogressors preferentially maintain highly functional HIV-specific CD8+ T cells. Blood 107, 4781–4789 (2006).

35 Ciuffreda, D. et al. Polyfunctional HCV-specific T-cell responses are associated with effective control of HCV replication. Eur. J. Immunol. 38, 2665–2677 (2008).

36 Precopio, M. L. et al. Immunization with vaccinia virus induces polyfunctional and phenotypically distinctive CD8(+) T cell responses. J. Exp. Med. 204, 1405–1416 (2007).

37 Park, S. H. et al. Successful vaccination induces multifunctional memory T-cell precursors associated with early control of hepatitis C virus. Gastroenterology 143, 1048-1060.e4 (2012).

